# Cell-Mediated Immune Response after COVID 19 Vaccination in Patients with Inflammatory Bowel Disease

**DOI:** 10.1101/2022.02.21.22271234

**Authors:** Freddy Caldera, Francis A. Farraye, Brian M. Necela, Davitte Cogen, Sumona Saha, Arnold Wald, Nader D. Daoud, Kelly Chun, Ian Grimes, Megan Lutz, Melanie D. Swift, Abinash Virk, Adil E. Bharucha, Tushar C. Patel, Gregory J. Gores, Saranya Chumsri, Mary S. Hayney, Keith L. Knutson

## Abstract

**Introduction:** Most patients with IBD mount an antibody response to mRNA COVID-19 vaccines, but few studies have evaluated the cell mediated immune response (CMIR).

**Methods:** We performed a prospective study (HERCULES) to evaluate CMIR among patients with IBD and healthy controls (HC) after completion of the primary series of mRNA COVID-19 vaccines.

**Results:** One hundred 158 patients with IBD and 20 HC were enrolled. The majority (89%) of IBD patients developed a CMIR which was not different than HC (94%, p=0.6667). There was no significant difference (p=0.5488) in CMIR response between those not immunosuppressed (median 255 Spike T cells/million PBMC, IQR 146, 958) and immunosuppressed (median 377, IQR 123, 1440). There was also no correlation between antibody responses and CMIR (p=0.5215)

**Discussion:** Most patients with IBD achieved CMIR to a COVID-19 vaccine. Future studies are needed evaluating sustained CMIR and clinical outcomes.

## Introduction

Two mRNA coronavirus disease 2019 (COVID-19) vaccines, mRNA-1273 (Moderna) and BNT162b2 (Pfizer-BioNTech), are highly effective in the general population.(1) Several studies have demonstrated that the majority of patients with inflammatory bowel disease (IBD) (95-99%) are able to achieve a measurable antibody response after the two-dose mRNA vaccine primary series, and 100% have a measurable antibody response after three doses.(2-5) Those who were less likely to seroconvert were older in age, received the BNT162b2 vaccine, and be on combination therapy with an anti-tumor necrosis factor alpha (TNF)-alpha inhibitor and an immunomodulator. (3-5)

The “<u>H</u>umo<u>R</u>al and <u>C</u>ell<u>UL</u>ar initial and <u>S</u>ustained immunogenicity in patients with IBD” (HERCULES) study found that after vaccination for COVID-19, antibody concentrations were lower in patients with IBD than in healthy controls (HC).(5) However, the clinical relevance of these differences is unknown. Vaccine induced cell-mediated immune response (CMIR) is an important component for protection against viruses such as SARS-CoV2, but a paucity of data in patients with IBD exists. The aim of this study was to evaluate the CMIR of COVID-19 vaccine patients with IBD.

## Methods

This was a two center, prospective, non-randomized study comprised of patients with IBD and HC in the HERCULES cohort.(5) Participants with IBD were enrolled at the University of Wisconsin-Madison (UW) and Mayo Clinic Florida (MAYOFL) and 20 HC were enrolled at MAYOFL. The inclusion criteria for patients with IBD on stable medication therapy has been previously described.(5) HC were eligible if they were not on immunosuppressive therapy and had documentation that they completed an mRNA vaccine series. Completion of an mRNA vaccines series was confirmed by review of the Wisconsin Immunization Registry for those recruited at UW and via electronic health records for those recruited at MAYOFL. Similar to other COVID-19 immunogenicity clinical trials, the humoral immune response and CMIR were measured at 28–35 days after the 2-dose mRNA series in patients with IBD and at approximately 30 days in HCs.

The primary outcome was the CMIR to mRNA COVID-19 vaccines in patients with IBD. The secondary outcomes were a comparison of the (1) CMIR in patients with IBD and HC and (2) CMIR and humoral immune response in all participants. (See Supplementary Methods for full details of measurement of humoral immune response, CMIR to SARS-CoV2 spike protein, and statistical analysis). The study received Institutional Review Board approval at the University of Wisconsin and Mayo Clinic Florida.

## Results

One hundred fifty-eight patients with IBD and 20 HC were enrolled. A greater proportion of HCs than patients with IBD (85% vs 54%) received the Pfizer vaccine (Table 1). The numbers of T cells responsive to spike antigens were evaluable in 151 patients with IBD and in 18 HC. The spike antibody levels were evaluable in 152 patients with IBD and in 18 HCs.

**Table 1.**
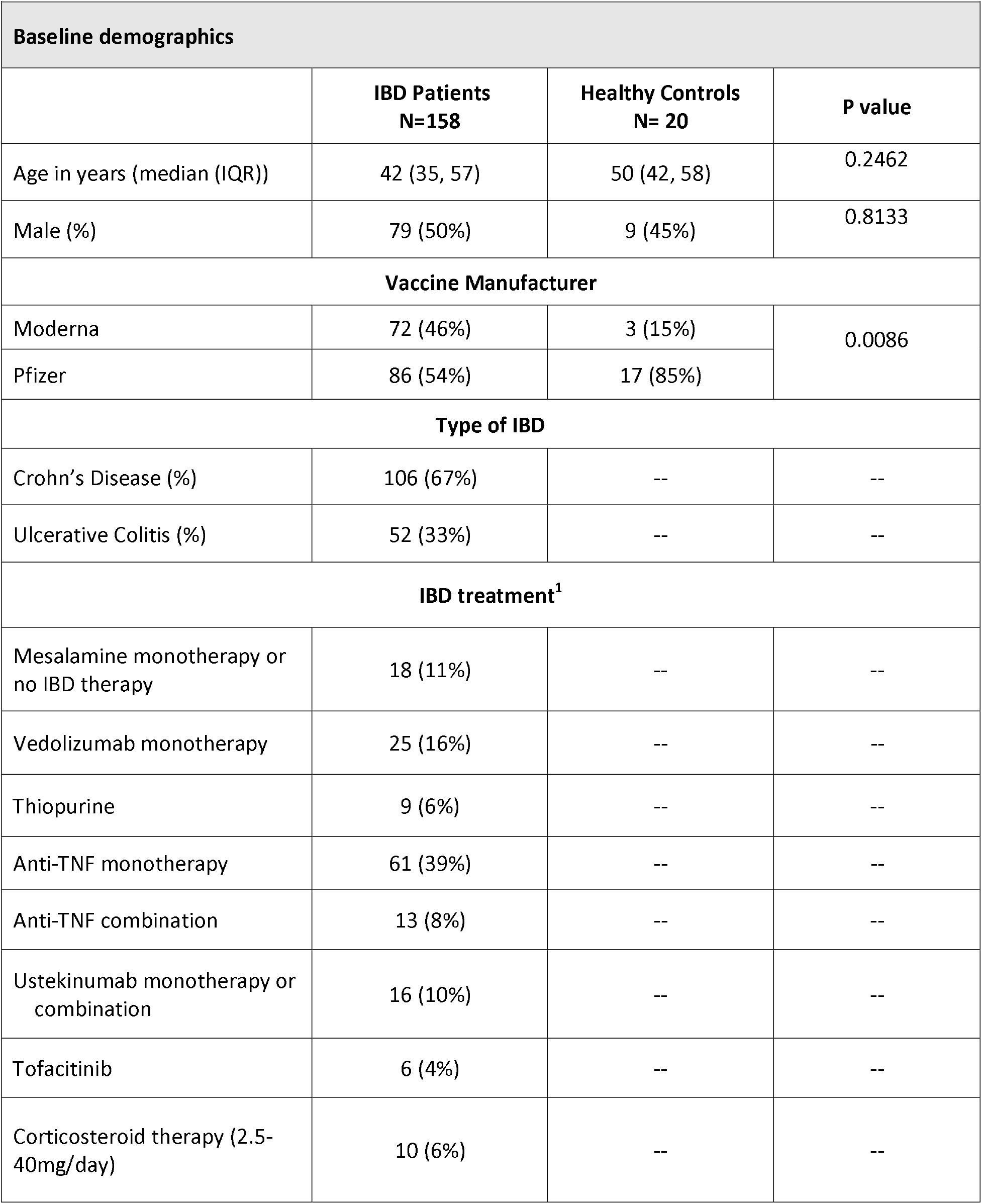
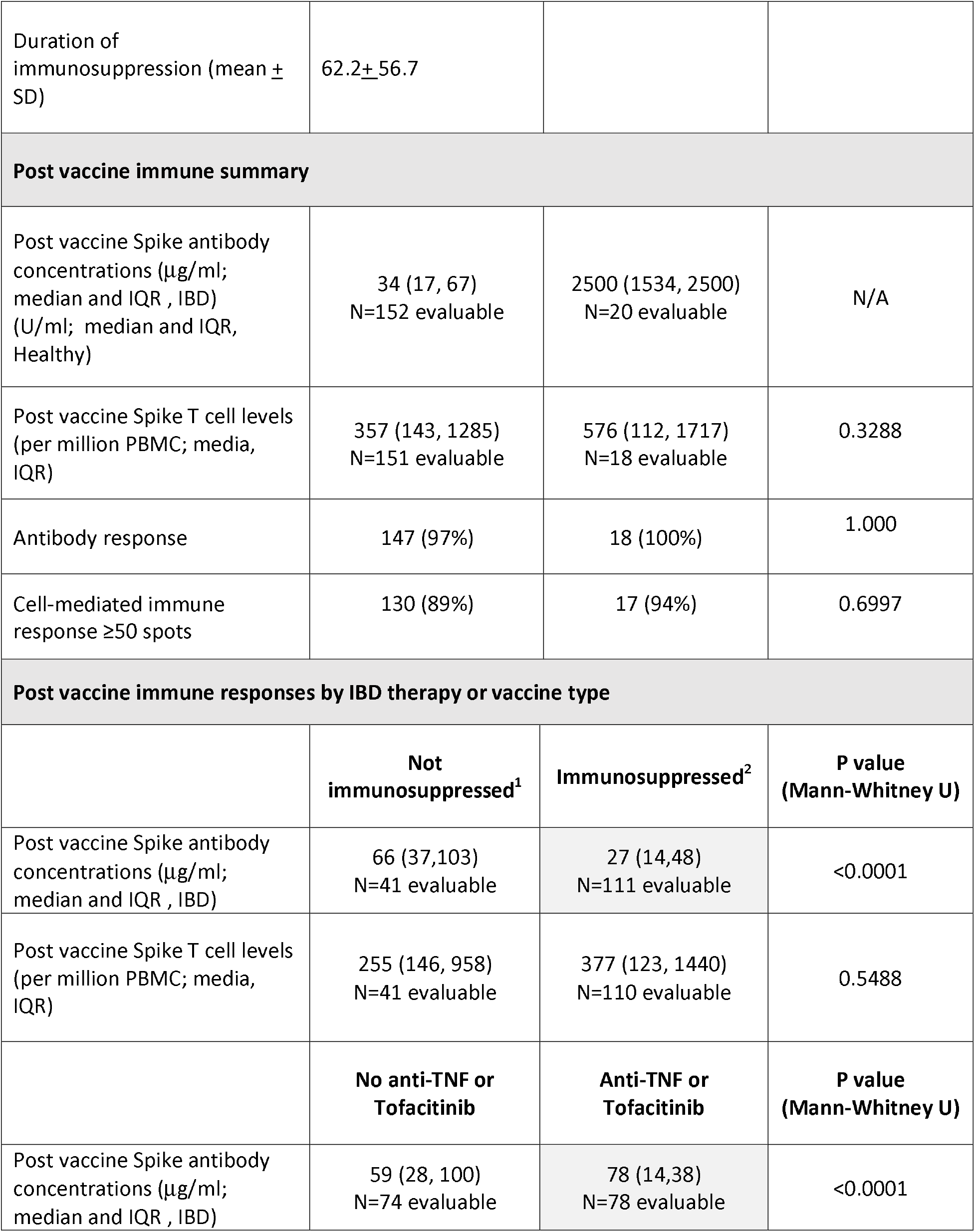
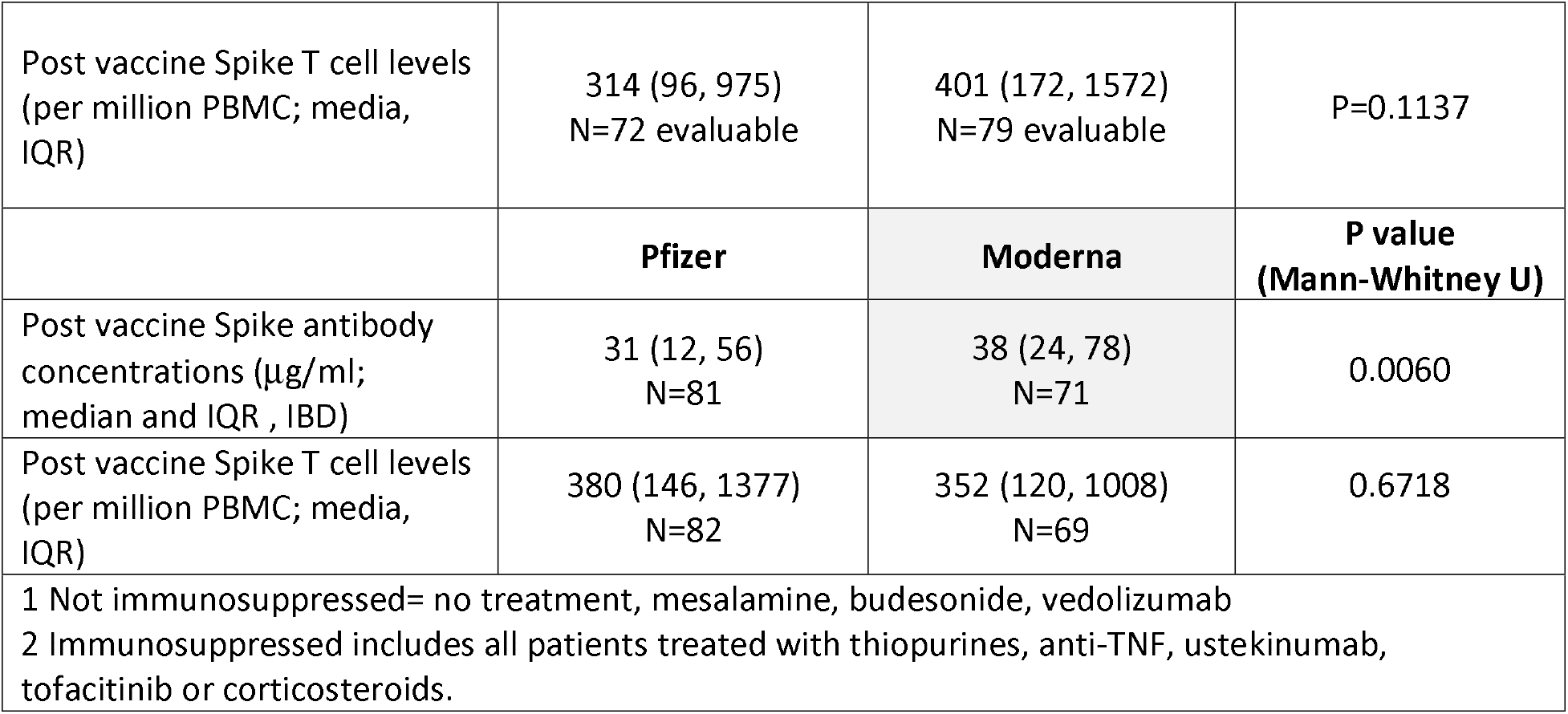

| Table 1 |  |  |  |
| --- | --- | --- | --- |
| Baseline demographics |  |  |  |
|  | IBD Patients<br>N=158 | Healthy Controls<br>N= 20 | P value |
| Age in years (median (IQR)) | 42 (35, 57) | 50 (42, 58) | 0.2462 |
| Male (%) | 79 (50%) | 9 (45%) | 0.8133 |
| Vaccine Manufacturer |  |  |  |
| Moderna | 72 (46%) | 3 (15%) | 0.0086 |
| Pfizer | 86 (54%) | 17 (85%) |  |
| Type of IBD |  |  |  |
| Crohn’s Disease (%) | 106 (67%) | -- | -- |
| Ulcerative Colitis (%) | 52 (33%) | -- | -- |
| IBD treatment<sup>1</sup> |  |  |  |
| Mesalamine monotherapy or no IBD therapy | 18 (11%) | -- | -- |
| Vedolizumab monotherapy | 25 (16%) | -- | -- |
| Thiopurine | 9 (6%) | -- | -- |
| Anti-TNF monotherapy | 61 (39%) | -- | -- |
| Anti-TNF combination | 13 (8%) | -- | -- |
| Ustekinumab monotherapy or combination | 16 (10%) | -- | -- |
| Tofacitinib | 6 (4%) | -- | -- |
| Corticosteroid therapy (2.5-40mg/day) | 10 (6%) | -- | -- |
| Duration of immunosuppression (mean ± SD) | 62.2±56.7 | | |
| **Post vaccine immune summary** |  |  |  |
| Post vaccine Spike antibody concentrations (μg/ml; median and IQR , IBD) (U/ml; median and IQR, Healthy) | 34 (17, 67)<br>N=152 evaluable | 2500 (1534, 2500)<br>N=20 evaluable | N/A |
| Post vaccine Spike T cell levels (per million PBMC; media, IQR) | 357 (143, 1285)<br>N=151 evaluable | 576 (112, 1717)<br>N=18 evaluable | 0.3288 |
| Antibody response | 147 (97%) | 18 (100%) | 1.000 |
| Cell-mediated immune response ≥50 spots | 130 (89%) | 17 (94%) | 0.6997 |
| **Post vaccine immune responses by IBD therapy or vaccine type** |  |  |  |
|  | **Not immunosuppressed<sup>1</sup>** | **Immunosuppressed<sup>2</sup>** | **P value (Mann-Whitney U)** |
| Post vaccine Spike antibody concentrations (μg/ml; median and IQR , IBD) | 66 (37,103)<br>N=41 evaluable | 27 (14,48)<br>N=111 evaluable | <0.0001 |
| Post vaccine Spike T cell levels (per million PBMC; media, IQR) | 255 (146, 958)<br>N=41 evaluable | 377 (123, 1440)<br>N=110 evaluable | 0.5488 |
|  | **No anti-TNF or Tofacitinib** | **Anti-TNF or Tofacitinib** | **P value (Mann-Whitney U)** |
| Post vaccine Spike antibody concentrations (μg/ml; median and IQR , IBD) | 59 (28, 100)<br>N=74 evaluable | 78 (14,38)<br>N=78 evaluable | <0.0001 |
| Post vaccine Spike T cell levels<br>(per million PBMC; media,<br>IQR) | 314 (96, 975)<br>N=72 evaluable | 401 (172, 1572)<br>N=79 evaluable | P=0.1137 |
|  | **Pfizer** | **Moderna** | **P value**<br>**(Mann-Whitney U)** |
| Post vaccine Spike antibody<br>concentrations (µg/ml;<br>median and IQR , IBD) | 31 (12, 56)<br>N=81 | 38 (24, 78)<br>N=71 | 0.0060 |
| Post vaccine Spike T cell levels<br>(per million PBMC; media,<br>IQR) | 380 (146, 1377)<br>N=82 | 352 (120, 1008)<br>N=69 | 0.6718 |
| 1 Not immunosuppressed= no treatment, mesalamine, budesonide, vedolizumab<br>2 Immunosuppressed includes all patients treated with thiopurines, anti-TNF, ustekinumab,<br>tofacitinib or corticosteroids. |  |  |  |

Among HC, all but one had a CMIR and all had a humoral immune response to an mRNA vaccine series. Likewise, most IBD patients had a CMIR (89%) and a humoral immune response (97%) (Table 1, Figure 1A). Three of four participants with no measurable antibody did have a CMIR (76, 232, & 4600 Spike T cells/million PBMC). There was no association between levels of antibody and CMIR (Figure 1B). Among patients with IBD, the humoral but not CMIR response was lower in patients taking vs not taking immunosuppressive medication(s) (Figures 1C-D). Additionally, no difference in Spike T cell responses was found between those on anti-TNF therapy or JAK inhibitors compared to other therapies (Table 1).

**Figure 1.**
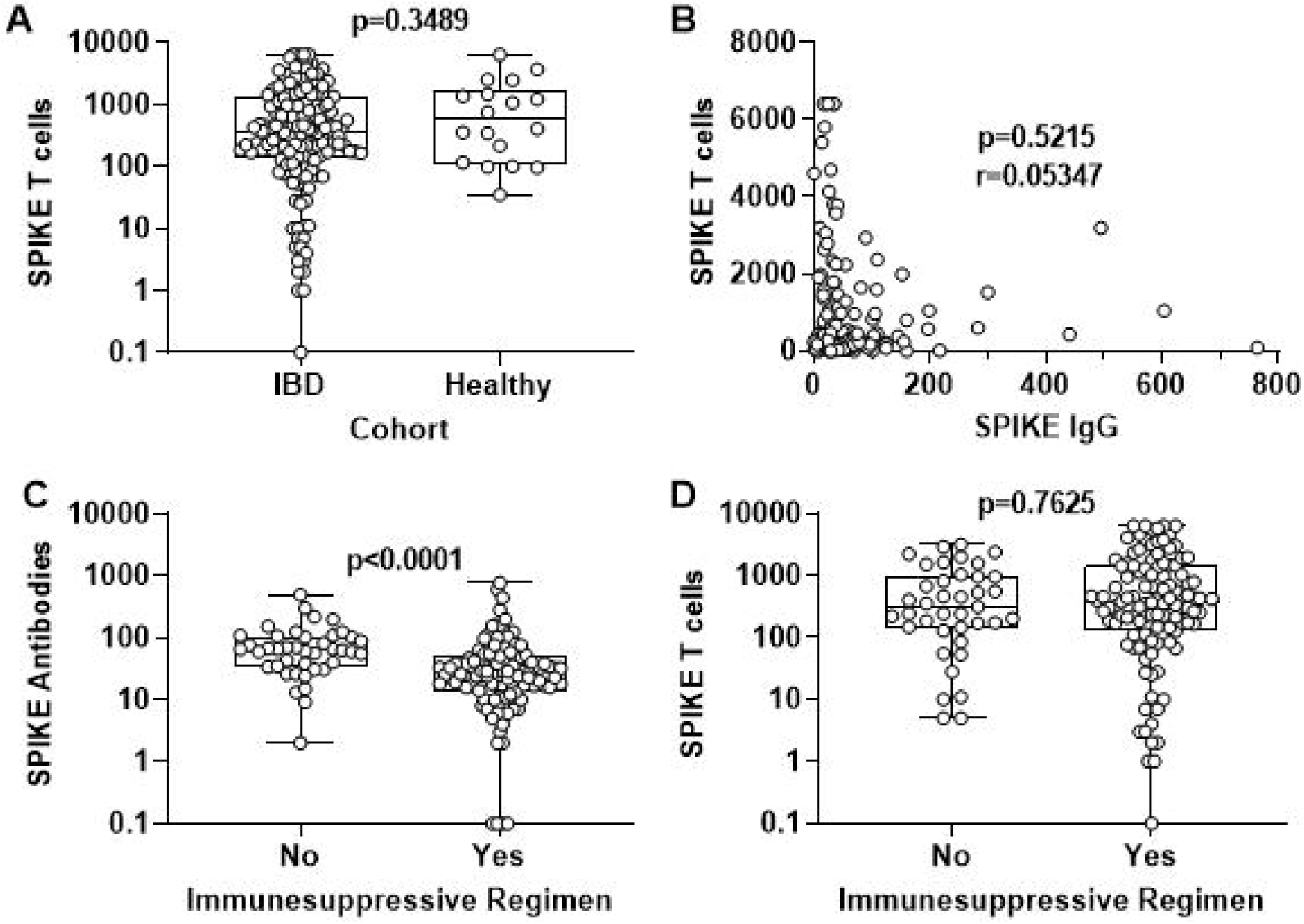
Humoral and cell mediated immune responses in IBD patients and normal healthy individuals following vaccination. ***A***, Box and whisker plot comparing Spike-specific T cell levels (per million PBMC) in all IBD patients and normal healthy controls. P value was calculated using the Mann-Whitney test. ***B***, Correlation plot comparing paired antibody (µg/ml serum) and Spike-specific T cell (per million PBMC) levels in patients with IBD. P value and r correlation coefficient were calculated using the Spearman correlation test. ***C-D***, Box and whisker plot comparing Spike-specific antibody levels (µg/ml serum) and Spike-specific T cell (per million PBMC) levels in IBD patients treated with either non-immunosuppressive or immunosuppressive regimens. P values were calculated using the Mann-Whitney test. Each symbol represents a unique patient or healthy donor.

## Discussion

In this study, essentially all patients with IBD, even patients on immunosuppressant medications, mounted a CMIR to the COVID-19 vaccine. By contrast to earlier studies, which observed lower antibody response after COVID-19 vaccination in immunosuppressed patients with IBD, the CMIR was not significantly different between those on immunosuppressants medications vs. those on non-immunosuppressive therapy in this study. Interestingly, we did not find a correlation between vaccine-induced antibody levels and CMIR.

Our findings contrast that seen in other immunosuppressed populations with CMIR rates of 36-46% in solid organ transplant recipients, 58% in those on B-cell depleting therapy, and 62-74% in patients with psoriasis on biological therapy and/or an immunomodulator. (6-9) Consistent with prior findings, humoral immune response studies in the above populations have shown that they have lower rates of antibody response after a primary mRNA series than those seen in this study. (7, 10)

Our study suggests that the humoral immune response provides an incomplete measure of COVID-19 vaccine-induced immune response. Additionally, our results and those of previous studies evaluating antibody responses to COVID-19 vaccines in patients with IBD suggest that most patients with IBD have a vaccine-induced immune response after an mRNA COVID-19 primary series similar to what has been seen in HC. This would suggest most patients with IBD may not require a third dose in the primary vaccine series. An additional dose to the primary series was recommended by the Advisory Committee on Immunization Practices (ACIP) for those who are moderately to severely immunocompromised. (11) This recommendation was largely based on evidence that solid organ transplant recipients who had a suboptimal antibody response (56%) after the primary series, and these data were extrapolated to other similarly immunosuppressed populations.(10) This additional dose to the primary series is intended for people who likely did not mount a protective immune response. A booster dose, preferentially an mRNA dose, is currently recommended for everyone age 12 years and older five months after their primary series for the general population and three months in moderately-severely immunosuppressed patients. Whether patients with IBD on immune-modifying therapy may benefit from an additional or earlier booster dose or a mix and match immunization strategy to reduce the incidence of breakthrough infections and/or severe disease is not known. It is unknown whether these therapies may impact sustained humoral and CMIR in patients with IBD.

Our study has several strengths. We evaluated patients on stable treatment regimens, and we used an established assay to measure CMIR, of which higher levels may be associated with better protection from disease. (12) We are limited by small samples in treatment groups and HC.

In summary, we found that almost all patients with IBD were able to mount CMIR which did not correlate with antibodies. Further studies are needed in evaluating sustained CMIR, the impact of booster dosing on CMIR, and long-term antibody concentrations and CMIR in patients with IBD.

## Data Availability

All data produced in the present study are available upon reasonable request to the authors

## Acknowledgements

The authors thank all the subjects who participated in the study, the specialty pharmacists at UW-Health for their help, and all the staff at the Office of Clinical Trials at University of Wisconsin-Madison for all their work in completion of the study. The authors acknowledge Edward Famularo at Mayo Florida, Sue McCrone, and Kayla Dillon, both at UW, for assisting with sample processing at Mayo Florida and Zhou Li for statistical advice.

## Supplementary Information

### Humoral immune response measurements

Nucleocapsid and Spike protein S1 receptor binding domain (RDB)-specific IgG antibodies were measured in sera at 28-35 days post completion of the two dose mRNA series in patients with IBD and at approximately 30 days in HC similarly to COVID-19 immunogenicity clinical trials. (13)

LabCorp’s Cov2Quant IgG assay uses electrochemiluminescence immunoassay technology for the quantitative measurement of IgG antibodies to SARS-CoV-2. This assay was used to measure anti-receptor binding domain IgG antibodies (the target of COVID-19 vaccines) and Roche anti nucleocapsid (indicative of a prior infection) antibodies in all patients with IBD. The Elecsys Anti-SARS-CoV-2 S electrochemiluminescence immunoassay (Roche Diagnostics, Switzerland) was used to measure the antibody responses in the normal healthy controls. Patients with prior COVID-19 infection (as assessed with an FDA-approved nucleocapsid antibody test) and patients on immunosuppressive therapy for an indication other than cancer were excluded. The sensitivity and correlation to neutralizing antibodies has been previously described. (5)

### Fluorospot Analysis

Fluorospot assays were performed to quantitate antigen-specific T cells capable of secreting interferon-γ (IFN-γ) with use of the human IFN-γ Fluorospot^Plus^ kit (Mabtech). Cryopreserved peripheral blood mononuclear cells (PBMCs) were thawed at 37°C, washed twice with RPMI media with 10% AB serum (Gemini Bio-Products), and viability determined by trypan blue exclusion using the Cellometer Vision (Nexcelom Bioscience). Only samples with > 85% viability were used in the assay. PMBCs were plated at 2.5⍰×⍰10^5^ per well in triplicate in 96-well round bottom plates and incubated at 37⍰°C, 5% CO^2^ for 24⍰hr. with complete medium alone, Spike protein peptide pools 1 + 2 (Stemcell Technologies, 1µg/ml), or phytohemagglutinin (PHA, 7.5 µg/ml, positive control). The SARS-CoV-2 Spike protein peptide pools 1 + 2 are pools of 158 peptides each consisted of 15-mer peptides with 11-amino acid overlaps that span amino acids 1-1273 of the spike protein. After 241’hours, cells were transferred to fluorospot plates pre-coated with anti-IFN-γ and that were blocked for 2 hr. with complete media at 37 °C. Plates were incubated for additional 24 hr, washed, and incubated with biotinylated anti-IFN-γ and streptavidin-550 conjugates with washes between each step. After the final wash, plates were incubated for 15 min with fluorescence enhancer-II, and after its removal, dried under a hood blower for 15 min. Plates were read on an AID ELISpot reader (San Diego, CA) using the Cy3 filter. AID Spot parameters were as follows: intensity (min 14; max 250); size min 43, max 5000); emphasis (small) and algorithm C. Antigen-specific T cells were defined as the average number of spots elicited by the antigen of interest minus the average number of spots elicited with culture medium alone. For each patient, the number of Spike-specific T cells was calculated by summing the individual responses to pools 1 and 2. For samples where spots were too numerous to count, spot number was set to 6400. All spot numbers were multiplied by four to achieve a standardized spots per million cells. Six patients with IBD and two healthy controls were excluded in the final analysis due to lack of PHA response. Although the lack of a PHA could indicate profound therapy-induced immune suppression, it could also indicate poor cell quality or lost sample, thus the results were not included. One IBD patient was excluded due to pre-vaccine positive COVID nucleocapsid response.

### Data analysis and statistical design

IBD treatment groups were defined as patients on stable doses of maintenance therapy in the following groups. The non-systemic immunosuppressive group: on mesalamine monotherapy or no therapy for IBD, or on vedolizumab monotherapy. Vedolizumab was considered in this group since previous studies have shown that it does not appear to impact vaccine response (14, 15).

Categorical variables were reported as frequencies (percentages) and continuous variables were reported as median with interquartile range. Mann-Whitney test was used to compare continuous variables between groups and Fisher’s exact test was used to compare categorical variables. Spearman’s test was used to evaluate for correlations between antibody and T cell responses. All tests were two sided with p value < 0.05 considered statistically significant.

